# *HLA* in isolated REM sleep behavior disorder and Lewy body dementia

**DOI:** 10.1101/2023.01.31.23284682

**Authors:** Eric Yu, Lynne Krohn, Jennifer A. Ruskey, Farnaz Asayesh, Dan Spiegelman, Zalak Shah, Ruth Chia, Isabelle Arnulf, Michele T.M. Hu, Jacques Y. Montplaisir, Jean-François Gagnon, Alex Desautels, Yves Dauvilliers, Gian Luigi Gigli, Mariarosaria Valente, Francesco Janes, Andrea Bernardini, Birgit Högl, Ambra Stefani, Abubaker Ibrahim, Anna Heidbreder, Karel Sonka, Petr Dusek, David Kemlink, Wolfgang Oertel, Annette Janzen, Giuseppe Plazzi, Elena Antelmi, Michela Figorilli, Monica Puligheddu, Brit Mollenhauer, Claudia Trenkwalder, Friederike Sixel-Döring, Valérie Cochen De Cock, Luigi Ferini-Strambi, Femke Dijkstra, Mineke Viaene, Beatriz Abril, Bradley F. Boeve, Guy A. Rouleau, Ronald B. Postuma, Sonja W. Scholz, the International LBD Genomics Consortium, Ziv Gan-Or

**Affiliations:** Department of Human Genetics, McGill University, Montréal, QC, Canada; The Neuro (Montreal Neurological Institute-Hospital), McGill University, Montréal, QC, Canada; Department of Neurology and Neurosurgery, McGill University, Montréal, QC, Canada; Neurodegenerative Diseases Research Unit, National Institute of Neurological Disorders and Stroke, Bethesda, MD, USA; Neuromuscular Diseases Research Section, National Institute on Aging, Bethesda, MD, USA; Sleep Disorders Unit, Pitié Salpêtrière Hospital, Paris Brain Institute and Sorbonne University, Paris, France; Oxford Parkinson’s Disease Centre (OPDC), University of Oxford, Oxford, United Kingdom; Division of Neurology, Nuffield Department of Clinical Neurosciences, University of Oxford, Oxford, United Kingdom; Center for Advanced Research in Sleep Medicine, Centre Intégré Universitaire de Santé et de Services Sociaux du Nord-de-l’Île-de-Montréal – Hôpital du Sacré-Coeur de Montréal, Montréal, QC, Canada; Department of Psychiatry, Université de Montréal, Montréal, QC, Canada; Department of Psychology, Université du Québec à Montréal, Montréal, QC, Canada; Department of Neurosciences, Université de Montréal, Montréal, QC, Canada; National Reference Center for Narcolepsy, Sleep Unit, Department of Neurology, Gui-de-Chauliac Hospital, CHU Montpellier, University of Montpellier, Inserm U1061, Montpellier, France; Clinical Neurology Unit, Department of Neurosciences, University Hospital of Udine, Udine, Italy; Department of Medicine (DAME), University of Udine, Udine, Italy; Sleep Disorders Clinic, Department of Neurology, Medical University of Innsbruck, Innsbruck, Austria; Department for Sleep Medicine and Neuromuscular disease, University Hospital Muenster, Muenster, Germany; Department of Neurology and Centre of Clinical Neuroscience, Charles University, First Faculty of Medicine and General University Hospital, Prague, Czech Republic; Department of Neurology, Philipps University, Marburg, Germany; Department of Biomedical, Metabolic and Neural Sciences, University of Modena and Reggio-Emilia, Modena, Italy; IRCCS, Institute of Neurological Sciences of Bologna, Bologna, Italy; Neurology Unit, Movement Disorders Division, Department of Neurosciences, Biomedicine and Movement Sciences, University of Verona, Verona, Italy; Department of Medical Sciences and Public Health, Sleep Disorder Research Center, University of Cagliari, Cagliari, Italy; Paracelsus-Elena-Klinik, Kassel, Germany; Department of Neurosurgery, University Medical Centre Göttingen, Göttingen, Germany; Sleep and Neurology Unit, Beau Soleil Clinic, Montpellier, France; EuroMov Digital Health in Motion, University of Montpellier IMT Mines Ales, Montpellier, France; Department of Neurological Sciences, Università Vita-Salute San Raffaele, Milan, Italy; Laboratory for Sleep Disorders, St. Dimpna Regional Hospital, Geel, Belgium; Department of Neurology, St. Dimpna Regional Hospital, Geel, Belgium; Department of Neurology, University Hospital Antwerp, Edegem, Antwerp, Belgium; Sleep disorder Unit, Carémeau Hospital, University Hospital of Nîmes, France; Department of Neurology, Mayo Clinic, Rochester, MN, USA; Department of Neurology, Johns Hopkins University Medical Center, Baltimore, MD, USA

## Abstract

**Background and Objectives:** Isolated/idiopathic REM sleep behavior disorder (iRBD) and Lewy body dementia (LBD) are synucleinopathies that have partial genetic overlap with Parkinson’s disease (PD). Previous studies have shown that neuroinflammation plays a substantial role in these disorders. In PD, specific residues of the human leukocyte antigen (*HLA*) were suggested to be associated with a protective effect. This study examined whether the *HLA* locus plays a similar role in iRBD, LBD and PD.

**Methods:** We performed HLA imputation on iRBD genotyping data (1,072 patients and 9,505 controls) and LBD whole-genome sequencing (2,604 patients and 4,032 controls) using the multi-ethnic HLA reference panel v2 from the Michigan Imputation Server. Using logistic regression, we tested the association of HLA alleles, amino acids and haplotypes with disease susceptibility. We included age, sex and the top 10 principal components as covariates. We also performed an omnibus test to examine which HLA residue positions explain the most variance.

**Results:** In iRBD, *HLA-DRB1*\*11:01 was the only allele passing FDR correction (OR=1.57, 95% CI=1.27-1.93, *p*=2.70e-05). We also discovered associations between iRBD and *HLA-DRB1* 70D (OR=1.26, 95%CI=1.12-1.41, *p*=8.76e-05), 70Q (OR=0.81, 95% CI=0.72-0.91, *p*=3.65e-04) and 71R (OR=1.21, 95% CI=1.08-1.35, *p*=1.35e-03). In *HLA-DRB1*, position 71 (*p*_omnibus_=0.00102) and 70 (*p*_omnibus_=0.00125) were associated with iRBD. We found no association in LBD.

**Discussion:** This study identified an association between *HLA-DRB1* 11:01 and iRBD, distinct from the previously reported association in PD. Therefore, the *HLA* locus may play different roles across synucleinopathies. Additional studies are required better to understand HLA’s role in iRBD and LBD.

## Introduction

Isolated/idiopathic REM sleep behavior disorder (iRBD) is a prodromal synucleinopathy characterized by enactment of dreams, vocalization and absence of muscle atonia during REM sleep.^1^ iRBD is one of the strongest predictors for certain neurodegenerative disorders, as approximately 80% of patients will convert to Parkinson’s disease (PD), Lewy body dementia (LBD) or multiple system atrophy (MSA) after 10-15 years on average following iRBD diagnosis.^2^

Previous evidence has shown that iRBD and synucleinopathies share a partial genetic overlap.^3^ While specific loci (*SNCA, GBA, TMEM175*) were shared between these traits, distinct loci such as *LRRK2* and *MAPT* for PD and *APOE* LBD were also identified.^3^ Furthermore, while the *SNCA* locus is important in PD, LBD and iRBD, the association with *SNCA* is driven by different variants for the different traits.^3^ Similar phenomenon occurs in the *SCARB2* locus, where different variants are associated with PD or RBD.^3^ Understanding the shared genes and pathways and the genetic differences will lead to better characterization of these disorders. For instance, microglial activation, a form of neuroinflammation, was found in all these disorders.^4-6^ However, the role of the immune system in their pathophysiology is poorly understood.

Recently, a fine-mapping study of the human leukocyte antigen (*HLA*) locus in PD demonstrated a strong association of HLA-DRB1 amino acids 11V, 13H and 33H with reduced PD risk.^7^ Located on chromosome 6, the *HLA* locus is a highly polymorphic region with complicated linkage patterns. *HLA* plays an essential role in the adaptive immune system by presenting antigens to T-cells.

Since the role of the *HLA* locus is unknown in iRBD and LBD, this study aims to examine whether *HLA* variants may affect the risk for these disorders. We analyzed the association of different *HLA* alleles, haplotypes and amino acids in two cohorts of iRBD and LBD patients.

## Methods

### Study population

iRBD and LBD cohorts from two previous genome-wide association studies (GWAS) were included in this analysis (Table 1).^3,8^ iRBD patients were diagnosed according to the International Classification of Sleep Disorders (2nd or 3rd Edition) with video polysomnography. LBD was diagnosed according to consensus criteria, as described elsewhere.^8-10^ The iRBD cohort is composed of 1,072 patients and 9,505 controls with genotyping data from the OmniExpress GWAS chip (Illumina inc.). The control group includes six publicly available cohorts: controls from the International Parkinson’s Disease Genomics Consortium (IPDGC) NeuroX dataset (dbGap phs000918.v1.p1), National Institute of Neurological Disorders and Stroke (NINDS) Genome-Wide genotyping in Parkinson’s Disease (dbGap phs000089.v4.p2), NeuroGenetics Research Consortium (NGRC) (dbGap phs000196.v3.p1), Parkinson’s Progression Markers Initiative (PPMI) and Vance (dbGap phs000394).

**Table 1:**
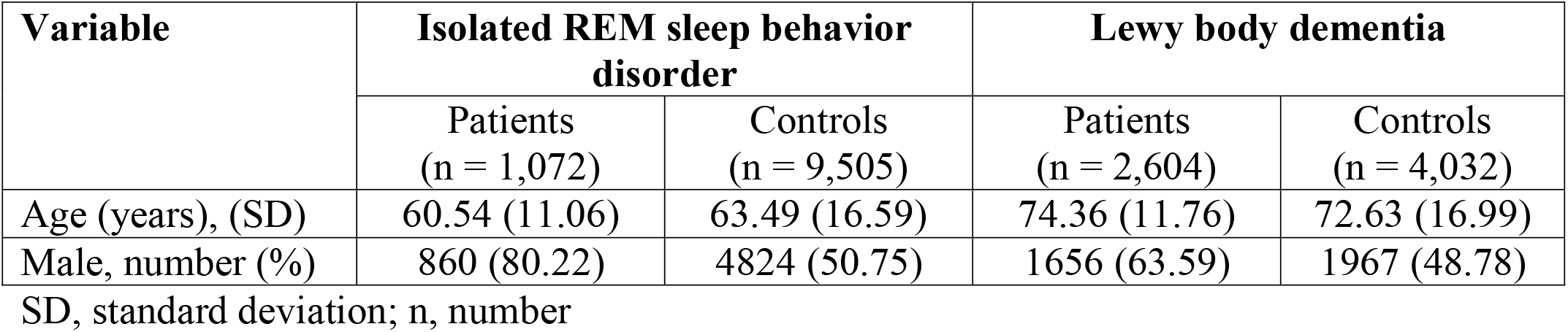
Study population after quality control.

| Variable | Isolated REM sleep behavior disorder |  | Lewy body dementia |  |
| --- | --- | --- | --- | --- |
|  | Patients<br>(n = 1,072) | Controls<br>(n = 9,505) | Patients<br>(n = 2,604) | Controls<br>(n = 4,032) |
| Age (years), (SD) | 60.54 (11.06) | 63.49 (16.59) | 74.36 (11.76) | 72.63 (16.99) |
| Male, number (%) | 860 (80.22) | 4824 (50.75) | 1656 (63.59) | 1967 (48.78) |

SD, standard deviation; n, number

The LBD cohort consisted of 2,604 patients and 4,032 controls with whole-genome sequencing data as described elsewhere.^8^ Study participants signed informed consent forms and the Institutional Review Board at McGill University approved the study protocol.

### Quality control

We performed standard GWAS quality control steps for both cohorts using PLINK v1.90. We excluded variants that were heterozygosity outliers (|F| > 0.15), sample call rate outliers (<0.95) and samples failing sex checks were also excluded. We determined genetic ancestry by merging samples with HapMap3 and clustering with principal components analysis (PCA). We only selected samples of European ancestry. A relatedness check was performed with GCTA^11^ to remove third-degree relatives or closer ones. Then, we performed several variant-level filtrations, such as removing call rate outliers (<0.95) and variants with significantly different missingness between cases and controls (p<0.0001). We also excluded variants that failed PLINK –test-mishap (p<0.0001) and deviated from Hardy-Weinberg equilibrium (p<0.0001) in controls.

### HLA imputation

Samples passing quality control were imputed on the Michigan Imputation Server with the four-digit multi-ethnic HLA reference panel v2^12^ using Minimac4 and phased with Eagle v2.4. This reference panel is composed of five global populations (n=20,349). Only alleles with imputation score (r^2^) above 0.8 were included. We determined *HLA* haplotypes using haplo.stats R package (https://analytictools.mayo.edu/research/haplo-stats/), which employs an Expectation– maximization (EM) algorithm.

### Power calculations

We performed power calculations online for each cohort using CaTS to compute statistical power. (https://csg.sph.umich.edu/abecasis/gas_power_calculator/). We assumed a prevalence of 1% for iRBD^13^ and 4% for LBD^14^. In iRBD, we had enough statistical power (>0.8) to detect an association (*p*=0.0005) with an odd ratio of 1.6 with a minor allele frequency (MAF) of 0.05. In LBD, we had enough statistical power (>0.8) to detect an association (*p*=0.0005) with an odd ratio of 1.4 with a MAF of 0.05.

### Statistical analysis

We performed logistic regression with an additive model on each *HLA* allele, haplotype and amino acid after adjusting for age at onset, sex and the top 10 principal components. We also performed an Omnibus test using the OMNIBUS_LOGISTIC module from HLA-TAPAS.^12^ All rare associations (carrier frequency < 1%) were excluded. A 5% false-discovery rate (FDR) for multiple testing was applied.

### Data availability

Anonymized data not published within this article will be made available by request from any qualified investigator.

### Code availability

All scripts used in this study can be found at https://github.com/gan-orlab/HLA_syn.

## Results

After *HLA* imputation, we examined the association of *HLA* alleles, haplotypes and amino acids. *HLA-DRB1*\*11:01 was the only allele passing FDR correction (OR=1.57, 95% CI=1.27-1.93, *p*=2.70e-05, Table 2). In addition, *HLA-DRB1* 70D, an amino acid present in *DRB1*\*11:01, was associated with iRBD (OR=1.26, 95%CI=1.12-1.41, *p*=8.76e-05). We also found association with 70Q (OR=0.81, 95% CI=0.72-0.91, *p*=3.65e-04 and 71R (OR=1.21, 95% CI=1.08-1.35, *p*=1.35e-03). In *HLA-DRB1*, positions 71 (*p*_omnibus_=0.00102) and 70 (*p*_omnibus_=0.00125) were the most associated with iRBD. *DRB1*\*11:01 also tags three haplotypes: *DQA1*\*05:01∼*DQB1*\*03:01∼*DRB1*\*11:01 (OR=1.40, 95%CI=1.16-1.70, *p*=5.17e-04), *DQA1*\*05:01∼*DRB1*\*11:01 (OR=1.41, 95%CI=1.16-1.72, *p*=5.43e-04), *DQB1*\*03:01∼*DRB1*\*11:01 (OR=1.36, 95%CI=1.13-1.64, *p*=1.04e-03).

**Table 2:**
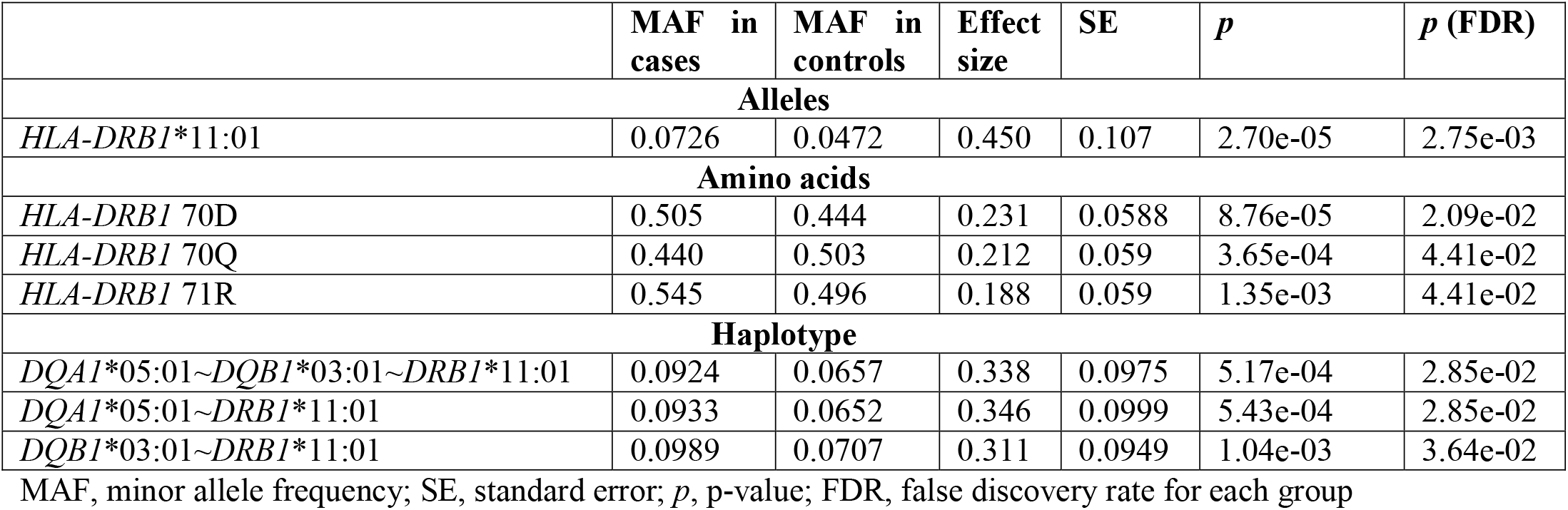
HLA association in isolated REM sleep behavior disorder.

When we repeated the analysis at one-field (two-digit) resolution, e.g., treating *DRB1*\*11:01 and 11:04 as the same, the association of *DRB1*\*11 was not significant (*p*=0.004, Supplementary Table #1), suggesting that it is specifically the *DRB1*\*11:01 allele associated with RBD. For LBD, no association was statistically significant after correction for multiple comparisons. We also examined the association of HLA-DRB1 33H, which was previously reported to be associated with PD (Supplementary Table #3).^7^ The MAFs of HLA-DRB1 33H in iRBD cases and controls were 0.125 vs. 0.149, respectively (*p*=0.499). Meanwhile, the DRB1 33H allele frequency in both LBD cases and its controls was 0.145.

## Discussion

This study shows an association between *DRB1*\*11:01, DRB1 70D, 70Q and 71R on iRBD. We also identified HLA-DRB1 positions 71 and 70 via an omnibus test, which suggests that residues at those positions explain a large amount of variance. HLA-DRB1 position 70-74 is a strong risk factor for rheumatoid arthritis and is referred to as a “shared epitope” (SE).^15^ The SE, in combination with DRB1 11V, was associated with a protective effect for PD.^16^ The SE is composed of a Q/R-K/R-RAA sequence with important antigen-binding grooves. However, 11:01 does not have the SE and there was no association between alleles with the SE (01:01, 01:02, 04:01, 04:04, 04:05, 04:08, 10:01)^16^ and iRBD. These findings indicate that the effects of position 70 and 71 may be independent of the SE. Additional studies examining the role of HLA-DRB1 in PD and iRBD will be necessary.

In addition, DRB1 33H, a variant also associated with PD, was not significantly associated with iRBD or LBD. However, the difference in carrier frequency between iRBD cases and controls for DRB1 33H, similar to that seen in PD, suggests that our study may lack the power to detect this association in iRBD. A recent study has suggested a shared mechanism between PD, AD, amyotrophic lateral sclerosis and HLA-DRB1*04, harboring the 33H amino acid change.^17^ This subtype was associated with decreased neurofibrillary tangles in post-mortem brains. It also binds to a K311 acetylated Tau PHF6 sequence.^17^ These results exemplify the possibility of different HLA types with specific genetic variants that may affect the binding of substrates relevant for neurodegenerative disorders and activating inflammatory response.

We could not replicate the association of a previous study of HLA antigens with 25 iRBD cases. This study showed a significant association between iRBD and DQB1*05 and DQB1*06.^18^ The most likely explanation for the discrepancy is that the previous study had reduced power to detect a true effect. Another study has suggested that HLA-DR expression was associated with iRBD.^19^ Fine-mapping and colocalization studies for these findings will be required once larger datasets of iRBD become available. Whether the mechanism underlying the associations with PD and iRBD is through functional effects of specific amino acid changes or due to different expressions of *HLA* genes in various brain tissues is still to be determined.

Although the role of the immune system in synucleinopathies is still unclear, some potential mechanisms of effect may exist. The varying effects of HLA between prodromal and clinical stages could be associated with HLA presenting different antigens in different brain regions. In LBD ^20^ and iRBD ^21^, activated CD4+ T-cells were shown to be dysregulated and associated with neuronal damage.

Another possibility is that the varying effects between iRBD and PD originate in the gastrointestinal tract.^22^ For example, constipation, a common symptom in the early stages of PD, can aggravate or be caused by gut inflammation. In iRBD patients, one study showed a prevalence of constipation between 18-41%.^22^ Gut bacterial antigens can be exposed from aging-related depletion of the gut lining. ^23^ HLA alleles may induce an immune response to self-proteins from these antigens.

Our study has several limitations. First, future replication studies with larger cohorts would be needed to increase statistical power since we do not have a replication cohort. Note that we used the most extensive available cohorts for iRBD and LBD.^3,8^ Due to the polygenicity of the HLA locus, various populations have different HLA allele frequencies. This study was done only on samples with European ancestry, and multi-ancestry analysis could provide more refined evidence on the role of HLA in synucleinopathies. The cohorts used in the study were also not matched for age and sex. However, we adjusted for these variables in the analysis.

To conclude, we found an alternative *HLA* association of iRBD compared to PD and LBD. More experimental evidence is necessary to characterize the genetic landscape of synucleinopathies and the role of the immune system.

## Acknowledgments

This work was financially supported by the Canadian Institutes of Health Research (#476751), the Michael J. Fox Foundation, Parkinson Canada, the Canadian Consortium on Neurodegeneration in Aging (CCNA), and the Canada First Research Excellence Fund (CFREF) awarded to McGill University for the Healthy Brains Healthy Lives (HBHL) program. The Montreal iRBD cohort is supported by the Canadian Institutes of Health Research. J.-F.G. holds a Canada Research Chair on Cognitive Decline in Pathological Aging. Z.G.O. is supported by the Fonds de recherche du QuebecSante (FRQS) Chercheurs boursiers award, and is a William Dawson and Killam Scholar. This research was supported in part by the Intramural Research Program of the U.S. National Institutes of Health (National Institute of NEurological Disorders and Stroke; project number: ZIANS003154). This work utilized the computational resources of the NIH HPC Biowulf cluster USA (http://hpc.nih.gov). We thank members of the International LBD Genomics Consortium (https://amp-pd.org/unified-cohorts/lbd#acknowledgements). A complete list of site investigators and acknowledgments for the International LBD Genomics Consortium is given in the Supplementary Information.

### Appendix I: Authors

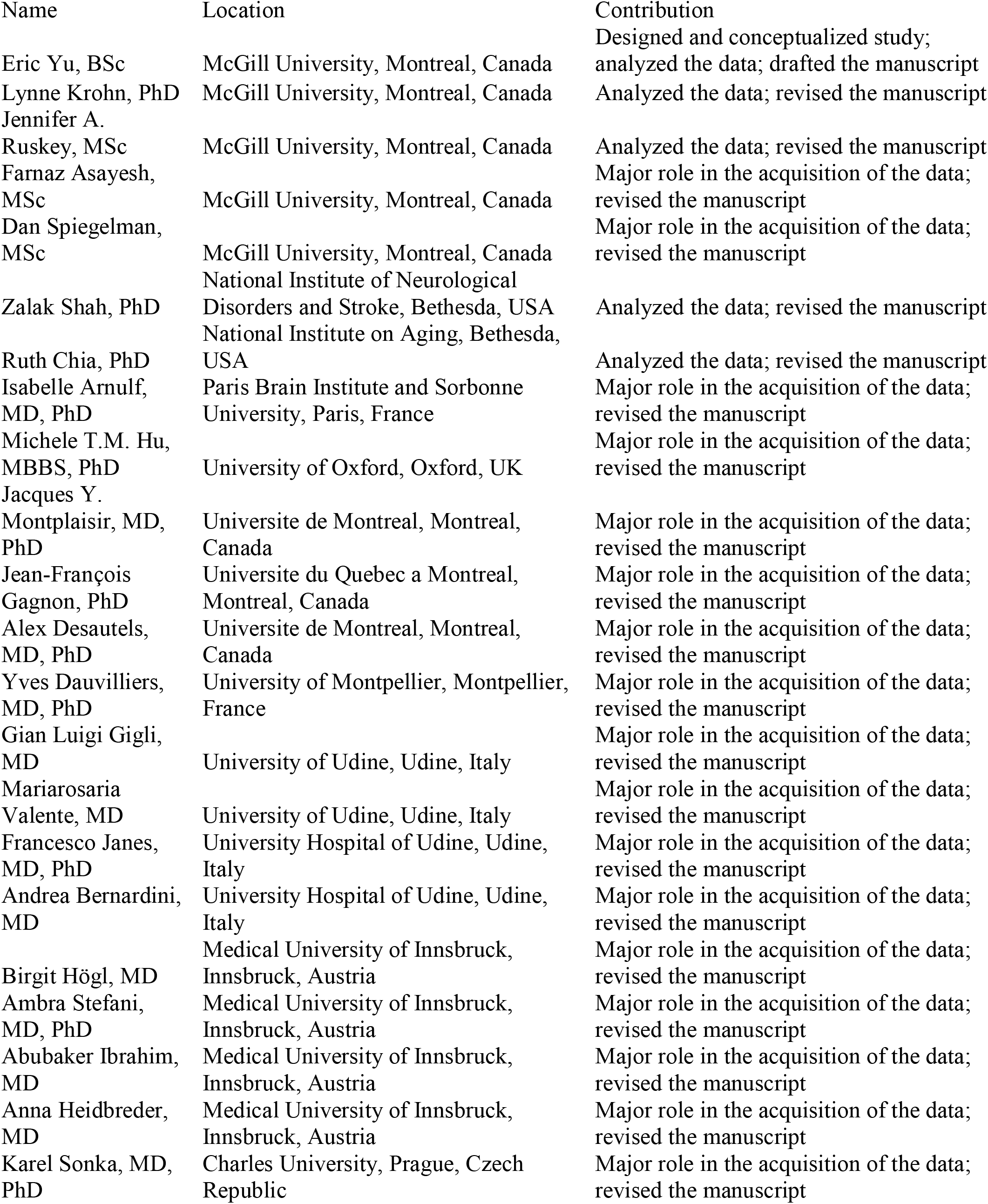
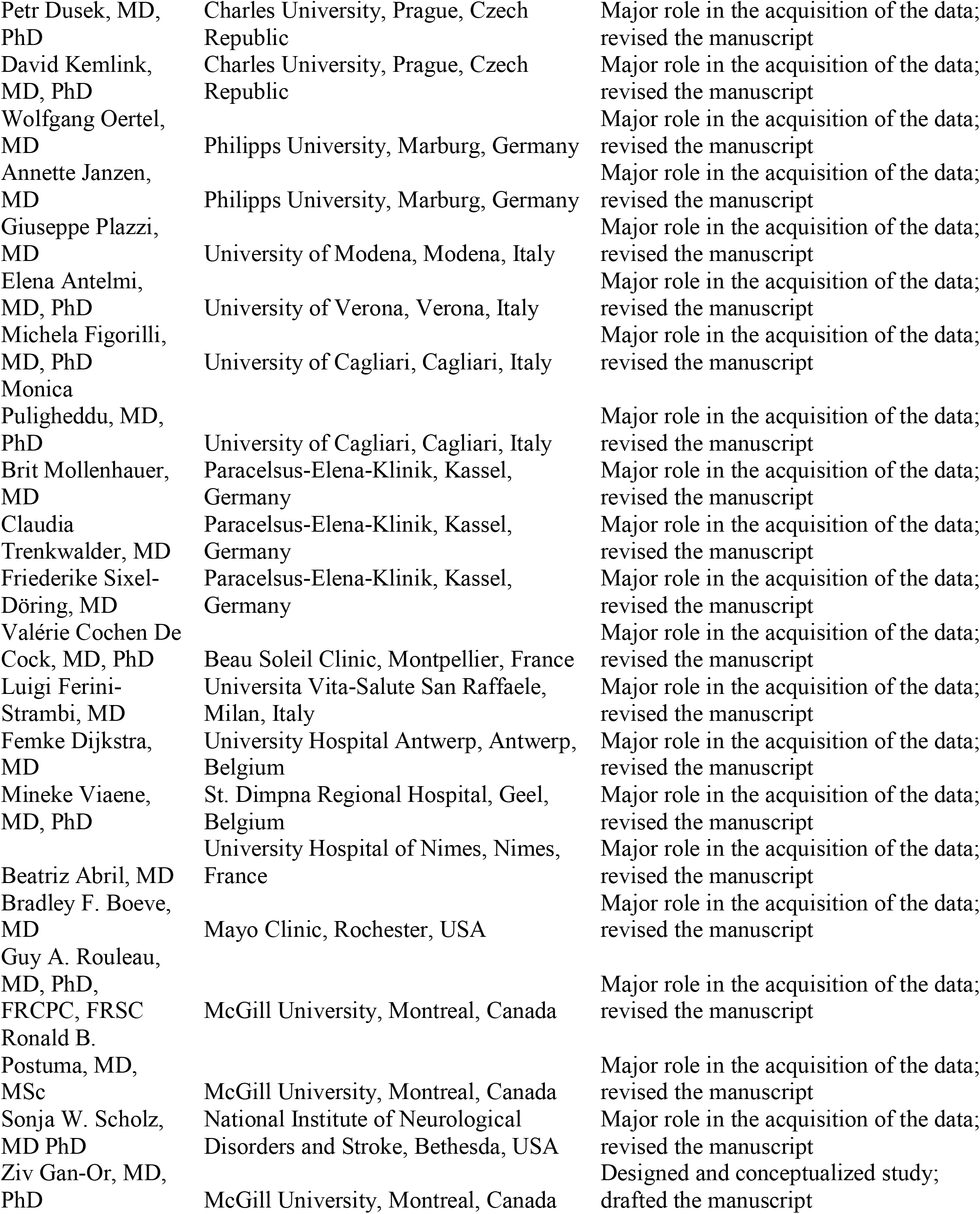

