## Supplemental Table 8 for "*HLA* in isolated REM sleep behavior disorder and Lewy body dementia"

**International LBD Genomics Consortium (iLBDGC)**

(Principal investigators for individual study sites are separated by country)

**Canada:** Sandra E. Black (Institute of Medical Science, Faculty of Medicine, University of Toronto, Toronto, ON, Canada; Division of Neurology, Department of Medicine, University of Toronto, Toronto, ON, Canada; Heart and Stroke Foundation Canadian Partnership for Stroke Recovery, Sunnybrook Health Sciences Centre, University of Toronto, Toronto, ON, Canada; Hurvitz Brain Sciences Research Program, Sunnybrook Research Institute, University of Toronto, Toronto, ON, Canada; LC Campbell Cognitive Neurology Research Unit, Sunnybrook Research Institute, University of Toronto, Toronto, ON, Canada), Ziv Gan-Or (Montreal Neurological Institute and Hospital, Department of Neurology & Neurosurgery, McGill University, Montreal, Canada), Julia Keith (Department of Anatomical Pathology, Sunnybrook Health Sciences Centre, University of Toronto, Toronto, ON, Canada), Mario Masellis (Cognitive & Movement Disorders Clinic, Sunnybrook Health Sciences Centre, University of Toronto, Toronto, ON, Canada; Division of Neurology, Department of Medicine, University of Toronto, Toronto, ON, Canada; Hurvitz Brain Sciences Research Program, Sunnybrook Research Institute, University of Toronto, Toronto, ON, Canada; LC Campbell Cognitive Neurology Research Unit, Sunnybrook Research Institute, University of Toronto, Toronto, ON, Canada), Ekaterina Rogaeva (Tanz Centre for Research in Neurodegenerative Diseases, University of Toronto, Toronto, ON, Canada).

**France:** Alexis Brice (Sorbonne Universites, Institute Du Cerveau – Paris Brain Institute, Paris, France), Suzanne Lesage (Sorbonne Universites, Institute Du Cerveau – Paris Brain Institute, Paris, France).

**Greece:** Georgia Xiromerisiou (Department of Neurology, University of Thessalia, University Hospital of Larissa, Larissa, Greece).

**Italy:** Andrea Calvo (“Rita Levi Montalcini” Department of Neuroscience, University of Turin, Turin, Italy), Antonio Canosa (“Rita Levi Montalcini” Department of Neuroscience, University of Turin, Turin, Italy), Adriano Chiò (“Rita Levi Montalcini” Department of Neuroscience, University of Turin, Turin, Italy; Institute of Cognitive Sciences and Technologies, C.N.R, Rome, Italy; Azienda Ospedaliero Universitaria Citta della Salute e della Scienza, Turin, Italy), Giancarlo Logroscino (Center for Neurodegenerative Diseases and the Aging Brain, University of Bari Aldo Moro At Pia Fondazione Panico Hospital-Tricase (LE), Bari, Italy), Gabriele Mora (ALS Center, Istituti Clinici Scientifici Maugeri, IRCCS Milano, Milan, Italy).

**Luxembourg:** Reijko Krüger (Luxembourg Center for Systems Biomedicine, University of Luxembourg, Esch-sur-Alzette, Luxembourg; Transversal Translational Medicine, Luxembourg Institute of Health, Strassen Luxembourg; Parkinson Research Clinic, Centre Hospitalier de Luxembourg, Luxembourg), Patrick May (Luxembourg Center for Systems Biomedicine, University of Luxembourg, Esch-sur-Alzette, Luxembourg).

**Spain:** Daniel Alcolea (Sant Pau Biomedical Institute, Hospital de la Santa Creu i Sant Pau, Universitat Autònoma de Barcelona, Barcelona, Spain; The Network Center for Biomedical Research in Neurodegenerative Diseases (CIBERNED), Madrid, Spain), Jordi Clarimon (Sant Pau Biomedical Research Institute, Hospital de la Santa Creu i Sant Pau, Universitat Autònoma de Barcelona, Barcelona, Spain; The Network Center for Biomedical Research in Neurodegenerative Diseases (CIBERNED), Madrid, Spain), Juan Fortea (Sant Pau Biomedical Research Institute, Hospital de la Santa Creu i Sant Pau, Universitat Autònoma de Barcelona, Barcelona, Spain; The Network Center for Biomedical Research in Neurodegenerative Diseases [CIBERNED], Madrid, Spain), Isabel Gonzalez-Aramburu (Institute for Research Marqués (IDIVAL), University of Cantabria and Department of Neurology, Marqués de Valdecilla Hospital, Santander, Spain; Centro de Investigación Biomédica en Red sobre Enfermedades Neurodegenerativas (CIBERNED), Madrid, Spain), Jon Infante (Institute for Research Marqués (IDIVAL), University of Cantabria and Department of Neurology, Marqués de Valdecilla Hospital, Santander, Spain; Centro de Investigación Biomédica en Red sobre Enfermedades Neurodegenerativas (CIBERNED), Madrid, Spain), Carmen Lage (Institute for Research Marqués (IDIVAL), University of Cantabria and Department of Neurology, Marqués de Valdecilla Hospital, Santander, Spain; Centro de Investigación Biomédica en Red sobre Enfermedades Neurodegenerativas (CIBERNED), Madrid, Spain), Alberto Lleó (Sant Pau Biomedical Research Institute, Hospital de la Santa Creu I Sant Pau, Universitat Autònoma de Barcelona, Barcelona, Spain; The Network Center for Biomedical Research in Neurodegenerative Diseases (CIBERNED), Madrid, Spain), Pau Pastor (Memory and Movement Disorders Units, Department of Neurology, University Hospital Mutua de Terrassa, Barcelona, Spain), Pascual Sanchez-Juan (Institute for Research Marqués (IDIVAL), University of Cantabria and Department of Neurology, Marqués de Valdecilla Hospital, Santander, Spain; Centro de Investigación Biomédica en Red sobre Enfermedades Neurodegenerativas (CIBERNED), Madrid, Spain).

**Republic of Ireland:** Francesca Brett (Dublin Brain Bank, Neuropathology Department, Beaumont Hospital, Dublin, Ireland).

**United Kingdom:** Dag Aarsland (Institute of Psychiatry, Psychology and Neuroscience [IoPPN], King’s College London, London, UK), Safa Al-Sarraj (Department of Clinical Neuropathology, King’s College Hospital and London Neurodegenerative Diseases Brain Bank, Institute of Psychiatry, Psychology and Neuroscience [IoPPN], King’s College London, London, UK), Johannes Attems (Translational and Clinical Research Institute, Campus for Ageing and Vitality, Newcastle University, Newcastle upon Tyne, UK), Steve Gentleman (Neuropathology Unit, Department of Brain Sciences, Imperial College London, London, UK), John A. Hardy (Department of Neurodegenerative Disease, UCL Queen Square Institute of Neurology, Queen Square, London, UK; UK Dementia Research Institute at University College London, UCL Institute of Neurology, University College London, London, UK; Reta Lila Weston Institute, UCL Queen Square Institute of Neurology, London, UK; UCL Movement Disorders Centre, University College London, London, UK), Angela K. Hodges (Institute of Psychiatry, Psychology and Neuroscience [IoPPN], King’s College London, London, UK), Seth Love (Dementia Research Group, School of Clinical Sciences, University of Bristol, Southmead Hospital, Bristol, UK), Ian G. McKeith (Translational and Clinical Research Institute, Campus for Ageing and Vitality, Newcastle University, Newcastle upon Tyne, UK), Christopher M. Morris (Translational and Clinical Research Institute, Campus for Ageing and Vitality, Newcastle University, Newcastle upon Tyne, UK), Huw R. Morris (Department of Clinical and Movement Neuroscience, UCL Queen Square Institute of Neurology, University College London, London, UK), Laura Palmer (South West Dementia Brain Bank, University of Bristol, Southmead Hospital, Bristol, UK), Stuart Pickering-Brown (Division of Neuroscience and Experimental Psychology, Faculty of Biology, Medicine and Health, The University of Manchester, Manchester, UK), Mina Ryten (NIHR Great Ormond Street Hospital Biomedical Research Centre, University College London, London, UK; Genetics and Genomic Medicine, Great Ormond Street Institute of Child Health, University College London, London, UK), Alan J. Thomas (Biomedical Research Building, Campus for Aging and Vitality, Newcastle University, Newcastle upon Tyne, UK), Claire Troakes (Institute of Psychiatry, Psychology and Neuroscience (IoPPN), King’s College London, London, UK).

**United States of America:** Marilyn S. Albert (Department of Neurology, Johns Hopkins University Medical Center, Baltimore, MD, USA), Matthew J. Barrett (Department of Neurology, University of Virginia School of Medicine, Charlottesville, VA, USA), Thomas G. Beach (Banner Sun Health Research Institute, Sun City, AZ, USA), Lynn M. Bekris (Genomic Medicine Institute, Cleveland Clinic, Cleveland, OH, USA), David A. Bennett (Rush Alzheimer’s Disease Center, Chicago, IL, USA), Bradley F. Boeve (Department of Neurology, Mayo Clinic, Rochester, MN, USA), Clifton L. Dalgard (Department of Anatomy, Physiology and Genetics, Uniformed Services University of the Health Sciences, Bethesda, MD, USA), Ted M. Dawson (Department of Neurology, Johns Hopkins University Medical Center, Baltimore, MD, USA; Neuroregeneration and Stem Cell Programs, Institute of Cell Engineering, Johns Hopkins University School of Medicine, Baltimore, MD, USA; Department of Pharmacology and Molecular Science, Johns Hopkins University School of Medicine, Baltimore, MD, USA; Solomon H. Snyder Department of Neuroscience, Johns Hopkins University School of Medicine, Baltimore, MD, USA), Dennis W. Dickson (Department of Neuroscience, Mayo Clinic, Jacksonville, FL, USA), Kelley Faber (Indiana University School of Medicine, Indianapolis, IN, USA), Tanis Ferman (Department of Psychiatry and Psychology, Mayo Clinic, Jacksonville, FL, USA), Margaret E. Flanagan (Northwestern University Feinberg School of Medicine, Chicago, IL, USA), Tatiana M. Foroud (Indiana University School of Medicine, Indianapolis, IN, USA), Bernardino Ghetti (Department of Pathology and Laboratory Medicine, Indiana University School of Medicine, Indianapolis, IN, USA), J. Raphael Gibbs (Laboratory of Neurogenetics, National Institute on Aging, MD, USA), Alison Goate (Ronald M. Loeb Center for Alzheimer’s disease, Nash Family Department of Neuroscience, Department of Genetics and Genomic Science, and Department of Pathology, Icahn School of Medicine at Mount Sinai, New York, NY, USA), David S. Goldstein (Clinical Neurocardiology Section, National Institute of Neurological Disorders and Stroke, Bethesda, MD, USA), Neill R. Graff-Radford (Department of Neurology, Mayo Clinic Florida, Jacksonville, FL, USA), Horacio Kaufmann (Department of Neurology, New York University School of Medicine, NY, USA), Walter A. Kukull (National Alzheimer’s Coordinating Center (NACC), University of Washington, Seattle, WA, USA), James B. Leverenz (Cleveland Lou Ruvo Center for Brain Health, Neurological Institute, Cleveland Clinic, OH, USA), Qinwen Mao (Northwestern University Feinberg School of Medicine, Chicago, IL, USA), Eliezer Masliah (Laboratory of Neurogenetics, National Institute on Aging, Bethesda, MD, USA), Edwin Monuki (University of California Irvine, Irvine, CA, USA), Kathy L. Newell (Department of Pathology and Laboratory Medicine, Indiana University School of Medicine, Indianapolis, IN, USA), Jose-Alberto Palma (Department of Neurology, New York University School of Medicine, NY, USA), Matthew Perkins (Department of Neurology, Michigan Medicine, University of Michigan, Ann Arbor, MI, USA), Olga Pletnikova (Department of Pathology [Neuropathology], Johns Hopkins University School of Medicine, Baltimore, MD, USA; Department of Pathology and Anatomical Sciences, Jacobs School of Medicine and Biomedical Sciences, University at Buffalo, Buffalo, NY, USA), Alan E. Renton (Ronal M. Loeb Center for Alzheimer’s disease and Nash Family Department of Neuroscience, Icahn School of Medicine at Mount Sinai, New York, NY, USA), Susan M. Resnick (Laboratory of Behavioral Neuroscience, National Institute on Aging, Baltimore, MD, USA), Liana S. Rosenthal (Department of Neurology, Johns Hopkins University Medical Center, Baltimore, MD, USA), Owen A. Ross (Department of Neuroscience, Mayo Clinic, Jacksonville, FL, USA; Department of Clinical Genomics, Mayo Clinic, Jacksonville, FL, USA), Clemens R. Scherzer (Harvard Medical School and Brigham & Women’s Hospital, Boston, MD, USA), Geidy E. Serrano (Banner Sun Health Research Institute, Sun City, AZ, USA), Vikram G. Shakkottai (Department of Neurology, University of Texas Southwestern Medical Center, Dallas, TX, USA), Ellen Sidransky (Medical Genetics Branch, National Human Genome Research Institute, Bethesda, MD, USA), Andrew B. Singleton (Laboratory of Neurogenetics, National Institute on Aging, Bethesda, MD, USA), Toshiko Tanaka (Longitudinal Studies Section, National Institute on Aging, Baltimore, MD, USA), Eric Topol (Scripps Research Translational Institute, Scripps Research, La Jolla, CA, USA), Ali Torkamani (Scripps Research Translational Institute, Scripps Research, La Jolla, CA, USA), Bryan J. Traynor (Laboratory of Neurogenetics, National Institute on Aging, Bethesda, MD, USA), Juan C. Troncoso (Department of Pathology [Neuropathology], Johns Hopkins University School of Medicine, Baltimore, MD, USA), Randy Woltjer (Department of Neurology, Oregon Health & Sciences University, Portland, OR, USA), Zbigniew K. Wszolek (Department of Neurology, Mayo Clinic Florida, Jacksonville, FL, USA), Sonja W. Scholz (Neurodegenerative Diseases Research Unit, National Institute of Neurological Disorder and Stroke, Bethesda, MD, USA; Department of Neurology, Johns Hopkins University Medical Center, Baltimore, MD, USA).

Acknowledgments for consortium members:

T.F. and K.F. report that samples provided by the National Centralized Repository for Alzheimer’s Diseases and Related Dementias (NCRAD) were supported under a cooperative agreement grant (U24 AG021886). This study used tissue samples and data that were provided by the Johns Hopkins Morris K. Udall Center of Excellence for Parkinson’s Disease Research (NIH P50 NS38377). ZKW is partially supported by the Mayo Clinic Center for Regenerative Medicine, Mayo Clinic in Florida Focused Research Team Program, the gifts from The Sol Goldman Charitable Trust, and the Donald G. and Jodi P. Heeringa Family, the Haworth Family Professorship in Neurodegenerative Diseases fund, and The Albertson Parkinson's Research Foundation. We are grateful to the Banner Sun Health Research Institute Brain and Body Donation Program of Sun City, Arizona, for the provision of human brain tissue and data. The Brain and Body Donation Program is supported by the National Institute of Neurological Disorders and Stroke (U24 NS072026 National Brain and Tissue Resource for Parkinson’s Disease and Related Disorders), the National Institute on Aging (P30 AG19610 Arizona Alzheimer’s Disease Core Center), the Arizona Department of Health Services (contract 211002, Arizona Alzheimer’s Research Center), the Arizona Biomedical Research Commission (contracts 4001, 0011, 05-901 and 1001 to the Arizona Parkinson’s Disease Consortium) and the Michael J. Fox Foundation for Parkinson’s Research. We are grateful to the Rush Alzheimer’s Disease Center for providing brain tissue and DNA samples, which was supported by the grants P30 AG10161, R01 AG15819, R01 AG17917, U01AG46152, U01 AG61356. We would like to thank the Canadian Consortium on Neurodegeneration in Aging (E. Rogaeva, Z. Gan-Or, M. Masellis). This research was supported in part by NIH grants P30 AG62677, U01 NS100620, U54 NS110435, The Mayo Clinic Dorothy and Henry T. Mangurian Jr. Lewy Body Dementia Program, the Little Family Foundation, and Ted Turner and Family LBD Functional Genomics Program.

Conflict of interest statement for consortium members:

ZKW serves as PI or Co-PI on Biohaven Pharmaceuticals, Inc. (BHV4157-206 and BHV3241-301), Neuraly, Inc. (NLY01-PD-1), and Vigil Neuroscience, Inc. (VGL101-01.001) grants. He serves as Co-PI of the Mayo Clinic APDA Center for Advanced Research and as an external advisory board member for the Vigil Neuroscience, Inc.
